# Role of pharmacist during the COVID-19 pandemic: a scoping review

**DOI:** 10.1101/2020.06.30.20143859

**Authors:** Marília Berlofa Visacri, Isabel Vitória Figueiredo, Tácio de Mendonça Lima

## Abstract

**Background:** Since the start of the new Coronavirus (COVID-19) outbreak in December 2019, pharmacists worldwide are playing a key role adopting innovative strategies to minimize the adverse impact of the pandemic.

**Objectives:** To identify and describe core services provided by the pharmacist during the COVID-19 pandemic.

**Methods:** A literature search was performed in MEDLINE, Embase, Scopus, and LILACS for studies published between December 1^st^, 2019 and May 20^th^, 2020 without language restriction. Studies that reported services provided by pharmacists during the COVID-19 pandemic were included. Two independent authors performed study selection and data extraction with a consensus process. The pharmacist’s intervention identified in the included studies were described based on key domains in the DEPICT v.2.

**Results:** A total of 1,189 records were identified, of which 11 studies fully met the eligibility criteria. Most of them were conducted in the United States of America (n=4) and China (n=4). The most common type of publication were letters (n=4) describing the workplace of the pharmacist in hospitals (n=8). These findings showed the different roles of pharmacists during the COVID-19 pandemic, such as disease prevention and infection control, adequate storage and drug supply, patient care and support for healthcare professionals. Pharmacists’ interventions were mostly conducted for healthcare professionals and patients (n=7), through one-to-one contact (n=11), telephone (n=6) or video conference (n=5). The pharmacists’ main responsibility was to provide drug information for healthcare professionals (n=7) as well as patient counseling (n=8).

**Conclusions:** A reasonable number of studies that described the role of the pharmacists during the COVID-19 pandemic were found. All studies reported actions taken by pharmacists, although without providing a satisfactory description. Thus, future research with more detailed description as well as an evaluation of the impact of pharmacist intervention is needed in order to guide future actions in this and-or other pandemic.

## Introduction

The Coronavirus Disease 2019 (COVID-19) is an infection caused by the Severe Acute Respiratory Syndrome Coronavirus 2 (SARS-CoV-2) first emerged in Wuhan (China) in December 2019, spreading rapidly across the world.^1^ On the 11^th^ of March 2020, the World Health Organization (WHO) declared COVID-19 a pandemic.^2^ At the time of writing there have been 10 million cases of COVID-19 reported globally, with more than 500 000 deaths reported across 216 countries.^3^ Currently, the COVID-19 pandemic is a major public health problem worldwide.

The most common symptoms for patients infected with COVID-19 are fever, cough, difficulty breathing, fatigue, and headache.^4^ Most symptomatic patients will develop mild symptoms. However, some patients may progress to serious illness, such pneumonia, acute respiratory distress syndrome, multi organ dysfunction and even death.^5^ So far, there are no proven effective treatments against COVID-19 and widespread effort is being devoted towards the development of a safe vaccine.^3^ Thus, the population must follow recommendations to decrease the transmission of SARS-CoV-2, including social distancing, wearing masks and strict hand hygiene.^6^

While millions of people are in their homes in order to decrease the risk of transmission of the infection, health workers are on the frontline against COVID-19.^7^ These professionals are committed to ensuring that the population have access to health services and to minimize the adverse impact of the pandemic. Given the seriousness of the coronavirus outbreaks, health professionals with expertise in public health are essential.

As healthcare professionals, pharmacists can play key role during the pandemic, acting directly with the community,^8^ continuing to care for patients with chronic diseases,^9,10^ working in hospital pharmacies and providing pharmaceutical care to COVID-19 patients.^11^ Moreover, they may provide reliable information for preventing, detecting, treating and managing coronavirus infections.^12,13^ As a result, several challenges have emerged and innovative strategies are being adopted by pharmacists to overcome them.^14^

Since the beginning of the outbreak, many guidelines have been published with recommendations for pharmacists as well as their responsibilities during the pandemic. However, few describe pharmacists’ experiences in this novel context. Therefore, this scoping review is aimed to identify and describe core services provided by the pharmacist during the COVID-19 pandemic.

## Method

A scoping review was performed to explore the literature, map and summarize the evidence regarding the role of the pharmacist during COVID-19 pandemic.^15^ This review was conducted following the recommendations of the Preferred Reporting Items for Systematic reviews and Meta-Analyses statement for Scoping Reviews (PRISMA-ScR)^16^ and the review protocol registered on Open Science Framework (https://doi.org/10.17605/OSF.IO/NE2GY).

### Search strategy

A comprehensive literature search was performed in the MEDLINE (PubMed), Embase, Scopus, and LILACS (Latin American and Caribbean Health Sciences Literature) databases published between December 1^st^, 2019 (first reports of COVID-19 in China) and May 20^th^, 2020 in order to identify relevant studies. The search strategy included combinations of terms relating the COVID-19 and pharmacy. The full strategies search for all databases can be found in S1 Appendix. Additionally, it was conducted a grey literature search in DOAJ - Directory of Open Access Journals (https://doaj.org/) aiming to identify not indexed studies in the databases listed above. No language restriction was applied. Duplicated studies were eliminated. In addition, references cited in all included articles were reviewed to identify any studies that might have been missed.

### Study selection

Studies that described services provided by the pharmacist during the COVID-19 pandemic were included. In addition, all publication types were eligible for inclusion. Studies that did not describe the role of the pharmacist during the COVID-19 pandemic; reviews, recommendations, and-or guidelines of pharmacist’s role during the pandemic; presented only pharmacotherapeutic options for COVID-19; and involved graduate students were excluded.

All titles and abstracts were independently screened and selected by the authors. Full-text articles were obtained and reviewed to determine whether the article met the eligibility criteria. If the full texts of the articles were not available in the databases, the corresponding authors were contacted by email or through ResearchGate (www.researchgate.net). Disagreements were resolved through discussion.

This process was performed using Rayyan QCRI, a free web application designed to help researchers working on systematic reviews.^17^

### Data extraction and analysis

For each included study, information such as the: author, date of publication or availability online, publication type, region, workplace of the pharmacist, participants, and results summary were extracted. The pharmacist interventions reported in these studies were described based on key domains in the Descriptive Elements of Pharmacist Intervention Characterization Tool (DEPICT) version 2: 1) contact with recipient (how the contact with the recipient occurs); 2) method of communication with recipient; 3) setting of the intervention (where the recipient received the service); 4) action(s) taken by pharmacist (what is done to address the identified problems); and 5) materials that support action(s) (items developed or provided by the pharmacist as part of the service).^18^ The authors independently completed the data extraction, using a preformatted spreadsheet in Microsoft Excel. Disagreements were resolved through discussion.

The results of this scoping review are presented as a narrative synthesis due to the heterogeneity of the studies included. The studies were categorized according to the characteristics of the publication and summarized in tables.

Following the PRISMA-ScR^16^, no quality assessment was performed due to the fact that scoping reviews aim to identify all the available evidence and highlight their main characteristics, regardless of the quality.

## Results

### Search results

The electronic search found 1,189 potentially relevant studies. After removing duplicates and reviewing the titles and abstracts, 62 articles were selected for full-text reading. In addition, no relevant studies were identified from searching the reference lists of the included studies. Of these, 11 studies met the inclusion criteria and were included for review. A flowchart of the literature search is shown in Fig. 1. The references for the excluded studies, with the reasons for their exclusion, are available in S2 Appendix.

**Figure 1.**
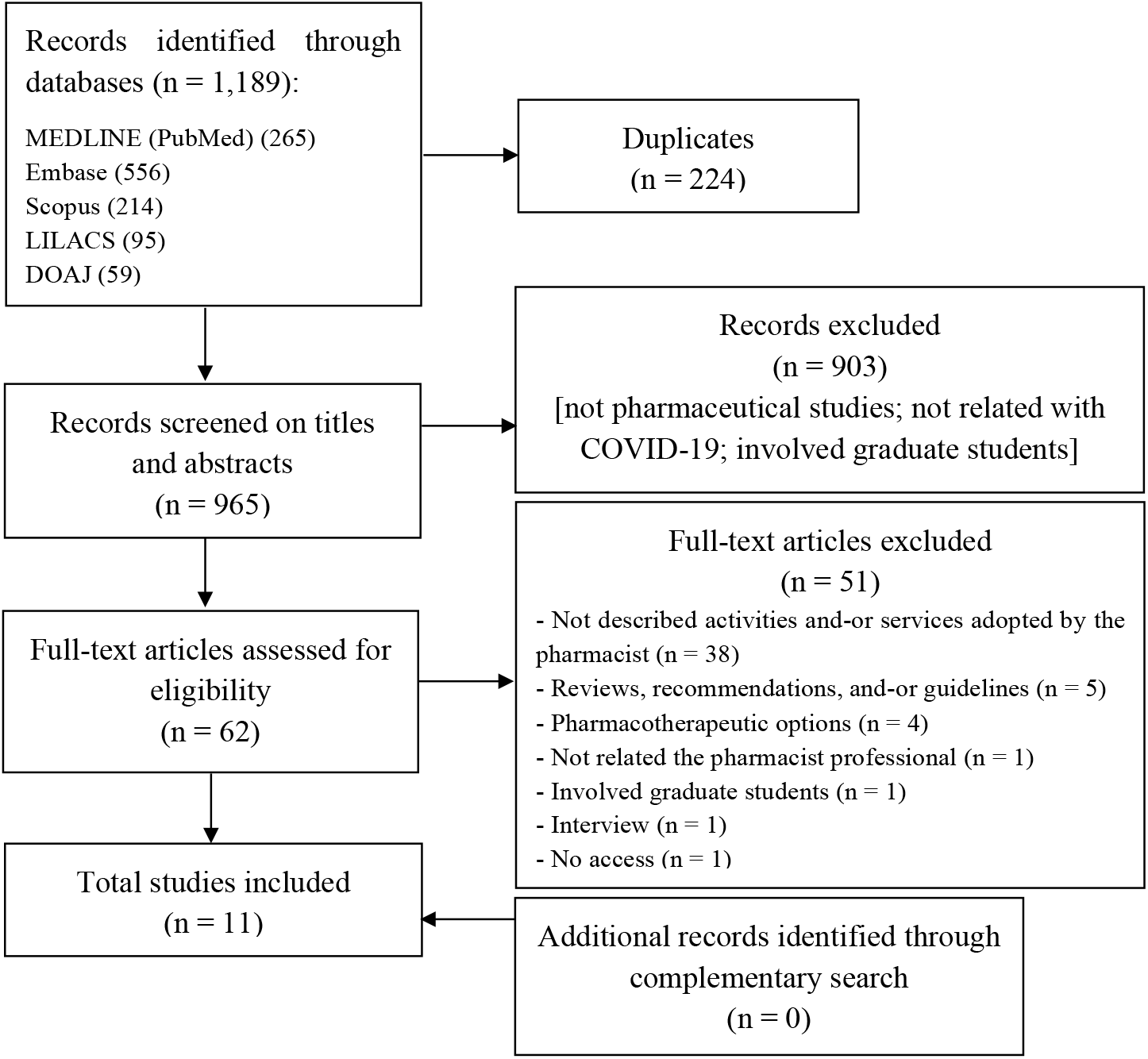
Study selection flowchart through literature search.

### Characteristics and summary of the results of the included studies

The characteristics of the 11 studies included in this scoping review are summarized in Table 1. Studies were conducted in the United States of America (n=4)^20,21,27,29^, China (n=4)^22,23,25,28^, Saudi Arabia (n=1)^19^, Taiwan (n=1)^24^, and Macao (n=1).^26^ All studies were published in English and reported between February and May 2020. The publication type of the included studies consisted of letters (n=4)^20,21,25,27^, research article (n=2)^19,22^, commentary (n=2)^23,26^, ideas and opinions (n=1)^24^, discussion (n=1)^28^, and note (n=1)^29^. The majority of the studies described the workplace of the pharmacist in hospitals (n=8)^19- 23,25,28,29^ following the ambulatory pharmacies (n=4)^19,20,28,29^, community pharmacies (n=2)^24,26^, and clinic (n=1).^27^ The participants of the included studies were miscellaneous, including the healthcare professionals (n=7)^19-23,28,29^, COVID-19 patients (n=5)^19,22,23,28,29^, general inpatient (n=2)^19,29^, general population (n=2)^24,26^, pediatric patients (n=1)^20^, solid organ transplant patients (n=1)^21^, patients on warfarin therapy (n=1)^25^, and myelofibrosis patients (n=1).^27^

**Table 1.** Characteristics of the included studies in the scoping review.

| **Author** | **Date of publication**<br>**(or available online)** | **Publication**<br>**Type** | **Region** | **Workplace of the**<br>**pharmacist** | **Participants** |
| --- | --- | --- | --- | --- | --- |
| Arain et al.<sup>19</sup> | 2020 May 18 | Research article | Saudi Arabia | Hospital and ambulatory pharmacy | Healthcare professionals and COVID-19 and non-COVID-19 patients |
| Elson et al.<sup>20</sup> | 2000 May 5 | Letter to the Editor | United States | Hospital and ambulatory pharmacy | Healthcare professionals and pediatric patients |
| Fan and Kamath<sup>21</sup> | 2020 May 4 | Letter to the Editor | United States | Hospital | Healthcare professionals and solid organ transplant patients |
| Hua et al.<sup>22</sup> | 2020 April 10 | Research article | China | Hospital | Healthcare professionals and COVID-19 patients |
| Meng et al.<sup>23</sup> | 2020 April 2 | Commentary | China | Hospital | Healthcare professionals and COVID-19 patients |
| Ou and Yang<sup>24</sup> | 2020 April 13 | Ideas and Opinions | Taiwan | Community pharmacy | Taiwan's population |
| Tan et al.<sup>25</sup> | 2020 May 13 | Letter | China | Hospital | Patients on warfarin therapy |
| Ung<sup>26</sup> | 2020 February 12 | Commentary | Macao | Community pharmacy | Macao's population |
| Yemm et al.<sup>27</sup> | 2020 May 14 | Letter to the<br>Editor | United States | Clinic | Myelofibrosis patients |
| Ying et al.<sup>28</sup> | 2020 April 6 | Discussion | China | Hospital<br>and ambulatory<br>pharmacy | Healthcare professionals<br>and COVID-19 patients |
| Zuckerman et<br>al.<sup>29</sup> | 2020 May 16 | Note | United States | Hospital and<br>ambulatory<br>pharmacy | Healthcare professionals<br>and COVID-19 and non-COVID-19 patients |

Abbreviation: COVID-19 (coronavirus disease 2019)

Table 2 shows a summary of the results of the included studies. All described actions taken by the pharmacist. However, only one study evaluated the outcomes associated with pharmacist intervention.^22^ Several services were related in these studies, including disease prevention and infection control^19,23,24,28^ (e.g., distribution of the masks, develop hygiene strategies, discard all unused drugs dispensed for COVID-19 patients, and social distancing), adequate storage and drug supply^19,20,22,23,28^ (e.g., drug formulary for treatment of COVID-19 to guide the drug supply and purchase, conversion of intravenous to oral medications when it was possible, and virtual communication for supply inventory), and patient care and support for healthcare professionals^19-23,25,27-29^ (e.g., ensure appropriate drug utilization for patients and healthcare professionals, participation of virtual rounds with interdisciplinary team, review online of the electronic orders, patient education, virtual medication consultation, and medication reconciliation).

**Table 2.**
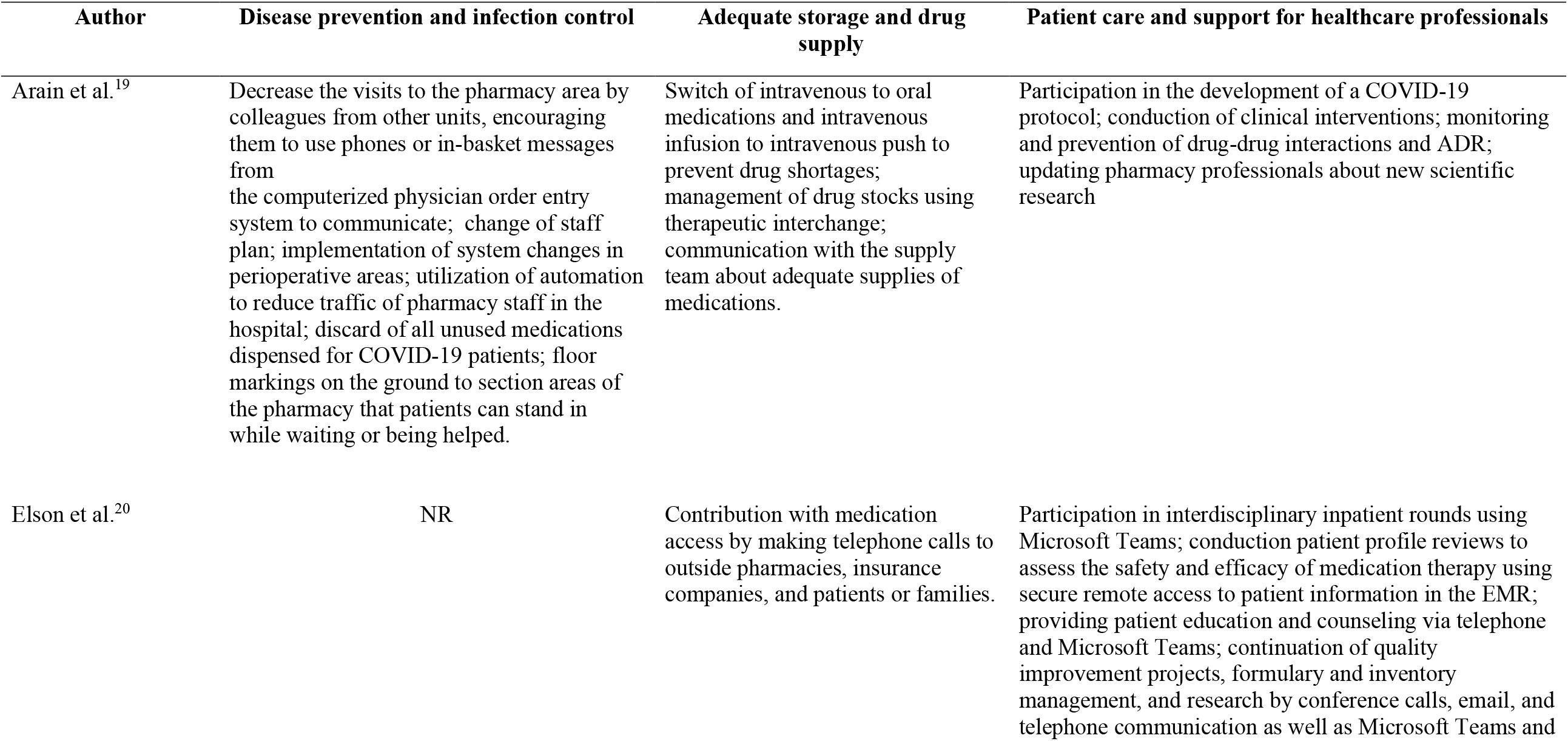
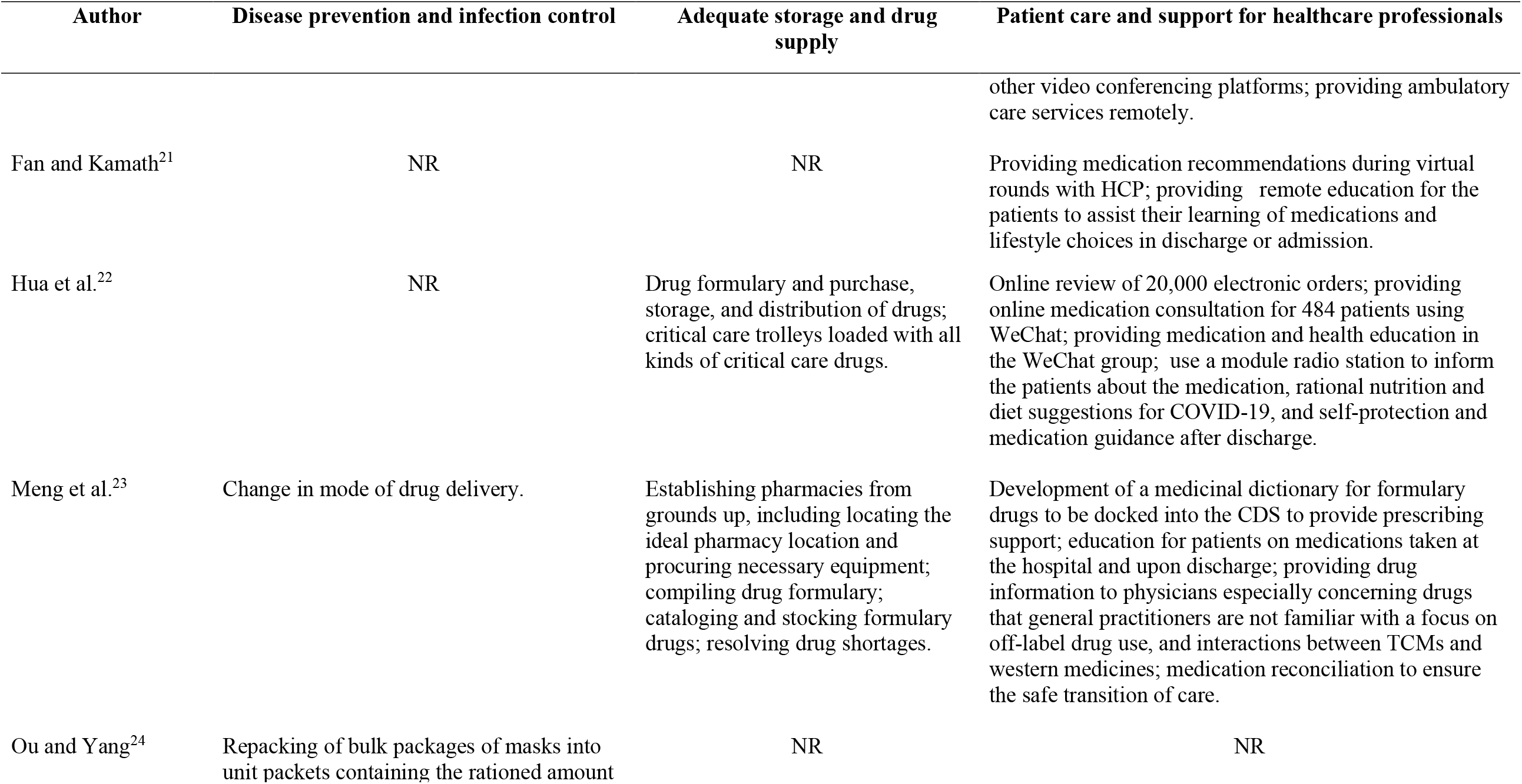
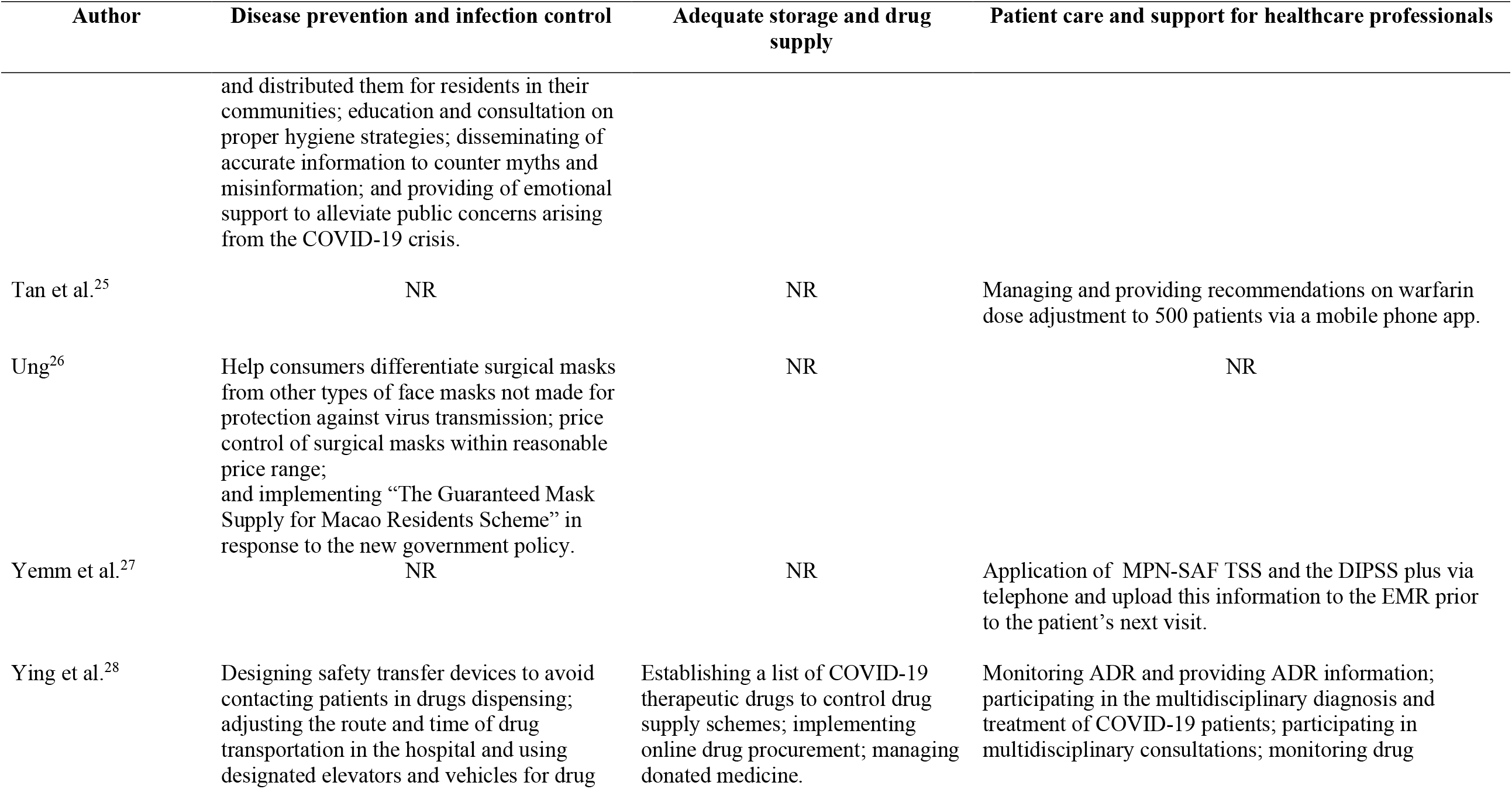
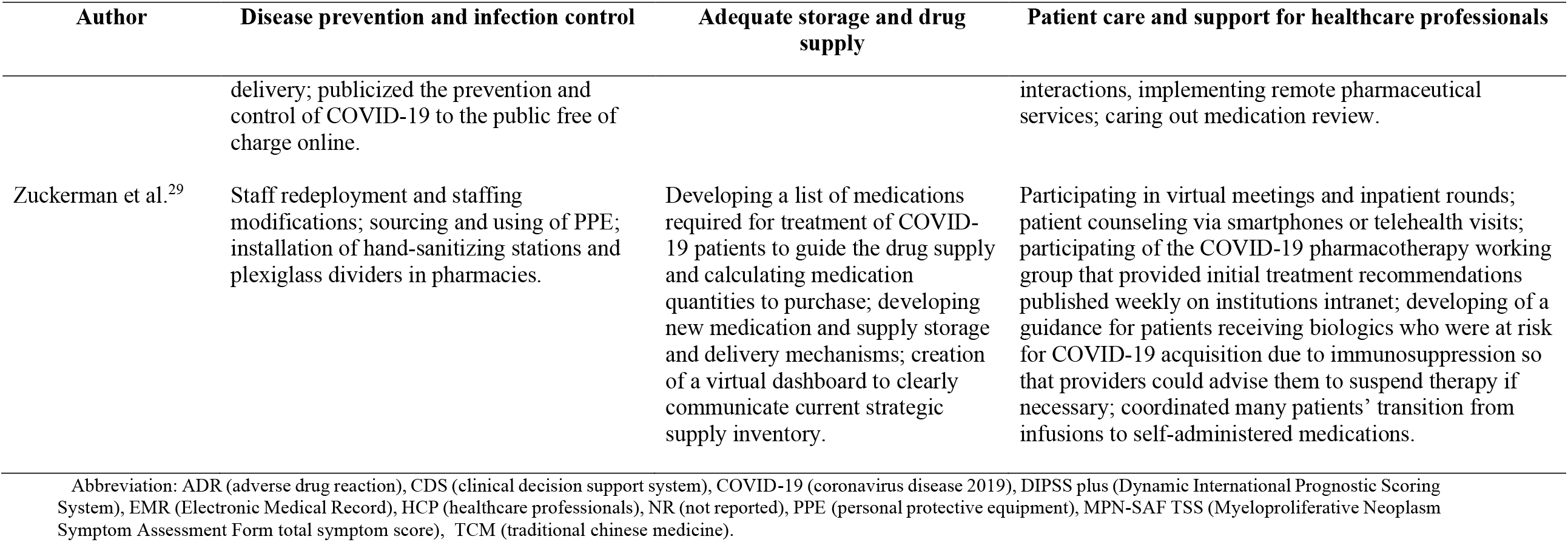
Summary of the results of the included studies in the scoping review.

### Characteristics of pharmacist interventions based on DEPICT 2

Studies were conducted for healthcare professionals and patients (n=7)^19-23,28,29^ or only patients (n=2).^25,27^ Two studies^24,26^ were conducted for the general population and, therefore, were classified as “not applicable”. All studies performed one-to-one contact with the recipients and six studies^19-22,28,29^ also used the group contact. Different methods of communication were reported, including face-to-face (n=4)^19,24,26,28^, written (n=5)^19,20,22,28,29^, telephone (n=6)^19,20,22,23,27,29^, video conference (n=5)^20-23,29^ and radio station.^22^ One study^25^ did not describe how the communication with the recipient has been performed. The studies were conducted at different setting of intervention, such as hospital bedside (n=7)^19-23,28,29^, hospital pharmacy (n=2)^19,29^, community pharmacy (n=2)^24,26^, ambulatory (n=4)^19,20,28,29^, and recipient’s home (n=5).^20,21,25,27,29^

Pharmacists had an important role in taking actions to address during the COVID-19 pandemic, including drug information for healthcare professionals (n=7)^19-23,28,29^, patient counseling (n=8)^19-23,25,28,29^, suggestion for change in therapy (n=3)^19,25,29^, monitoring results report (n=1)^19^, drug supply management (n=6)^19,20,22,23,28,29^, safety measures for infection control (n=4)^19,24,26,29^, and application of tools to evaluated a disease (n=1).^27^ Regarding the materials that support actions adopted by pharmacists, most studies (n=7)^20-22,24-27^ did not report them. Among those that described support resources provided by the pharmacists, educational materials (n=4)^19,23,28,29^ were the most reported, following the protocols (n=2)^19,29^, discharge letter (n=1)^19^, and safety alert system (n=1).^19^ The description of the pharmacist interventions according to DEPICT version 2 are shown in Table 3.

**Table 3.** Description of the pharmacist interventions according to the DEPICT version 2.

| **Author** | **Recipient** | **Contact with recipient** | **Methods of communication** | **Setting of the intervention** | **Action(s) taken by pharmacist** | **Materials that support action(s)** |
| --- | --- | --- | --- | --- | --- | --- |
| Arain et al.<sup>19</sup> | Patient and HCP | One-to-one (patient and HCP) and group (HCP) | Face-to-face (patient and HCP), written (patient and HCP), and telephone (HCP) | Hospital bedside (patient and HCP), hospital pharmacy (HCP), and ambulatory setting (patient) | Drug information for HCP; patient counseling; suggestion for change in therapy; monitoring results report; drug supply management; safety measures for infection control | Discharge letter; educational materials; protocol; safety alert system |
| Elson et al.<sup>20</sup> | Patient and HCP | One-to-one (patient) and group (HCP) | Written (patient and HCP), telephone (patient and HCP), and video conference (patient and HCP) | Hospital bedside (patients and HCP), ambulatory setting (patient), and recipient's home (patient) | Drug information for HCP; patient counseling; drug supply management | NR |
| Fan and Kamath<sup>21</sup> | Patient and HCP | One-to-one (patient) and group (HCP) | Video conference (patient and HCP) | Hospital bedside (patients and HCP) and recipient's home (patient) | Drug information for HCP; patient counseling | NR |
| Hua et al.<sup>22</sup> | Patient and HCP | One-to-one (patient and HCP) and group (patient) | Written (patient and HCP), telephone (patient), video conference (patient), and radio station (patient) | Hospital bedside (patient and HCP) | Drug information for HCP; patient counseling; drug supply management | NR |
| Meng et al.<sup>23</sup> | Patient and HCP | One-to-one (patient and HCP) | Telephone and video conference (patient and HCP) | Hospital bedside (patient and HCP) | Drug information for HCP; patient counseling; drug supply management; safety measures for infection control | Educational materials |
| Ou and Yang<sup>24</sup> | NA* | One-to-one | Face-to-face | Community pharmacy | Safety measures for infection control | NR |
| Tan et al.<sup>25</sup> | Patient | One-to-one | NR | Recipient's home | Patient counseling; suggestion for change in therapy | NR |
| Ung<sup>26</sup> | NA* | One-to-one | Face-to-face | Community pharmacy | Safety measures for infection control | NR |
| Yemm et al.<sup>27</sup> | Patient | One-to-one | Telephone | Recipient's home | Application of tools to evaluated a disease | NR |

| Author | Recipient | Contact with recipient | Methods of communication | Setting of the intervention | Action(s) taken by pharmacist | Materials that support action(s) |
| --- | --- | --- | --- | --- | --- | --- |
| Ying et al.<sup>28</sup> | Patient and HPC | One-to-one (patient and HPC) and group (HPC) | Face-to-face, written (patient and HPC) | Hospital bedside (patient and HCP) and ambulatory setting (patient and HCP) | Drug information for HPC; patient counseling; drug supply management; safety measures for infection control | Educational materials |
| Zuckerman et al.<sup>29</sup> | Patient and HCP | One-to-one (patient and HCP) and group (HCP) | Written (patients and HCP), telephone (patients), and video conference (HCP) | Hospital bedside (patient and HCP), hospital pharmacy (HCP), ambulatory setting (patient and HCP) and recipient's home (patient) | Drug information for HPC; patient counseling; suggestion for change in therapy; drug supply management; safety measures for infection control | Educational materials; protocols |

\*Pharmacist interventions were conducted for the general population.
Abbreviation: HCP (healthcare professionals), NA (not applicable), NR (not reported).

## Discussion

### Summary of evidence

This scoping review identified 11 relevant studies on the services provided by the pharmacist during the COVID-19 pandemic. To the best of our knowledge, this is the first review to discuss this question. These results indicate that there are a reasonable number of studies on this topic in a short time of the COVID-19 pandemic, although most of them are letters to the editor and other rapid scientific communications. Moreover, most studies were conducted in the United States and Asia, particularly in the region of China. These findings were expected because this region was the first local affected by the SARS-CoV-2 virus. Diversely, to our surprise, no studies were performed in Europe, especially in the United Kingdom, Italy and Spain, where the COVID-19 pandemic spread quickly in mid-March. It is important to note that most studies reported the hospital as the workplace of pharmacists. Experiences with community pharmacists should be encouraged since most of the time the community pharmacy is a first point-of-care of the patients.

In light of these findings, researchers must be engaged to design and report future studies with greater methodological rigor and more detailed description of the pharmacist interventions, in order to support and guide the actions of the pharmacists in this and-or other pandemic.

### General view of the studies

Most of the studies found in this review were conducted in the United States of America and China. These countries are the second and first in the scientific publication ranking worldwide, respectively.^30^ Moreover, China was the first region affected by the COVID-19 infection^1^ and therefore first experiences were felt in this location.

Regarding the type of publication, most studies were letters and other rapid scientific communications reporting experiences. These publications did not contain details of the work experiences and are at the lowest level of evidence.^31^ However, considering the current pandemic, when there is a need for rapid information for activity guidelines by the pharmacists need to have quick information to guide their activities, these publications are convenient and acceptable.

The hospital was the main workplace of the pharmacist in the included studies, which was an expected result because the role of pharmacist in hospital pharmacy practice is one of the most consolidated.^32^ Pharmacists have a very comprehensive role within the hospital, performing from administrative activities to clinical services.^33^ Therefore, they must be involved with all aspects of medicines use and be accessible as a point of contact for patients and health care providers.^34^ As a consequence, it is more than expected that they would be on the frontline against COVID-19 pandemic and reporting on their successful experiences.

This review showed that pharmacists may play an important role during the COVID-19 pandemic. The results reported were categorized as “disease prevention and infection control”, “adequate storage and drug supply” and “patient care and support for healthcare professionals”. These categories are basically the responsibilities that International Pharmaceutical Federation (FIP) has stated that it would like pharmacists to have in both primary care context (i.e. community pharmacies and primary healthcare facilities) and in hospital settings.^12^ Other recommendations of the scientific societies are also available and can help to direct the pharmacist intervention.^35-37^

According to the key domains of pharmacist interventions, most studies were carried out for healthcare professionals and patients. Pharmacist-provided interventions with the recipients have been shown to improve patient outcomes and contribute to substantial healthcare savings.^38^ In contrast, two studies did not classify as to the recipient of the intervention because the DEPICT version 2 did not predict interventions in the general population. It is important to note that pharmacists can play a role in health education and disease prevention in various ways for the general population.^39-42^ Moreover, the level of pharmacist-recipient interaction varied between studies. Most of them described the use of telephone, written interaction including web-based, and video conference. Studies involving the use of these methods of communication have been successful^43-45^ and these valuable tools may be applied in a social distancing context. In addition, one study reported that pharmacists used a hospital’s radio station as a communication strategy in patients who had difficulty in dealing with available technologies, highlighting the creative character.^22^ Regarding the setting of intervention, most studies were performed in hospital bedside, ambulatory, and recipient’s home. Pharmacist interventions were described in several settings^46-48^ as to improving the quality of care. Unfortunately, only two studies related the community pharmacy as the setting of intervention. A recent study discussed the role of community pharmacists during the COVID-19 pandemic, collecting and summarizing the experience of Chinese community pharmacies.^49^ It is well known that community pharmacies are an important setting of care in the COVID-19 pandemic period and further studies in this context should be encouraged.

Drug information for healthcare professionals and patient counseling were the main actions provided by the pharmacists identified in this review, similarly with other studies.^48,50^ These actions focus on enhancing the problem solving skills of the patient for the purpose of improving or maintaining the quality of life.^18^ In addition, other actions (drug supply management and safety measures for infection control) were also identified in some studies. Gross and MacDougall^51^ described these actions (e.g. planning the drug shortages and antiviral stewardship) as vital during the COVID-19 pandemic period. Moreover, the key domain “Materials that support actions” has not been reported in most studies. Among those that described these resources, most of them used educational materials. These findings are similar to a study that describes the role and impact of pharmacists in Spain.^52^ These resources are useful to support the pharmacy services.^18^ The lack of this information affected the understanding which tools were used by pharmacists in their actions.

### Limitations

This study has some limitations. It is possible that some studies were missed due to not being indexed in the databases searched or being published in websites of institutions or scientific societies. Moreover, the number of publications on COVID-19 is rapidly increasing in a short time and some studies of interest available after the established search period have not been included. Finally, this review did not analyze the quality of the studies taking into account the inherent characteristic of the scoping reviews.

## Conclusion

A reasonable number of studies that described the role of the pharmacists during the COVID-19 pandemic were found. Several methods of communication were performed in different settings of intervention. Moreover, all studies reported actions taken by pharmacists, mainly drug information and patient counseling, although description was not satisfactory. Thus, future research with more detailed description and evaluated the impact of pharmacist intervention is needed in order to guide the actions of the pharmacists in this and-or other pandemic.

## Data Availability

Yes

## Authors’ Contributions

MBV and TML identified the reports in the databases and collected data of the studies included. MBV and TML drafted the manuscript. IVF revised for finalization of the manuscript. All authors approved the final manuscript.

## Supplemental Files

Appendix 1. Search strategies of the scoping review

Appendix 2. List of excluded studies

## Appendix 1.

Search strategies of the scoping review

### A. MEDLINE (PubMed)

#1 “COVID-19” [Supplementary Concept] OR (2019 novel coronavirus disease) OR (COVID19) OR (COVID-19 pandemic) OR (SARS-CoV-2 infection) OR (COVID-19 virus disease) OR (2019 novel coronavirus infection) OR (2019-nCoV infection) OR (coronavirus disease 2019) OR (coronavirus disease-19) OR (2019-nCoV disease) OR (COVID-19 virus infection)

#2 “Pharmacists”[Mesh] OR (Pharmacist) OR (Clinical Pharmacists) OR (Clinical Pharmacist) OR (Pharmacist, Clinical) OR (Pharmacists, Clinical) OR (Community Pharmacists) OR (Community Pharmacist) OR (Pharmacist, Community) OR (Pharmacists, Community) OR (Hospital Pharmacists) OR “Pharmacies”[Mesh] OR (Community Pharmacy) OR (Community Pharmacies) OR “Pharmacy Service, Hospital”[Mesh] OR (Pharmaceutical Service, Hospital) OR (Service, Hospital Pharmaceutical) OR (Hospital Pharmacy Services) OR (Pharmacy Services, Hospital) OR (Services, Hospital Pharmacy) OR (Service, Hospital Pharmaceutic) OR (Hospital Pharmacy Service) OR (Hospital Pharmaceutic Service) OR (Hospital Pharmaceutic Services) OR (Pharmaceutic Services, Hospital) OR (Services, Hospital Pharmaceutic) OR (Pharmaceutic Service, Hospital) OR (Hospital Pharmaceutical Service) OR (Hospital Pharmaceutical Services) OR (Pharmaceutical Services, Hospital) OR (Services, Hospital Pharmaceutical) OR (Service, Hospital Pharmacy) OR (Pharmacy Service, Clinical) OR (Service, Clinical Pharmacy) OR (Clinical Pharmacy Services) OR (Pharmacy Services, Clinical) OR (Services, Clinical Pharmacy) OR (Clinical Pharmacy Service) OR “Pharmaceutical Services”[Mesh] OR (Services, Pharmaceutic) OR (Services, Pharmacy) OR (Pharmaceutic Services) OR (Pharmaceutic Service) OR (Service, Pharmaceutic) OR (Services, Pharmaceutical) OR (Pharmaceutical Service) OR (Service, Pharmaceutical) OR (Pharmacy Services) OR (Pharmacy Service) OR (Service, Pharmacy) OR (Pharmaceutical Care) OR (Care, Pharmaceutical) OR “Medication Therapy Management”[Mesh] OR (Management, Medication Therapy) OR (Therapy Management, Medication) OR (Drug Therapy Management) OR (Management, Drug Therapy) OR (Therapy Management, Drug)

#1 AND #2

Filtrer: publication date from 2019/12/01.

### B. Embase

#1 ‘covid 19’/exp OR ‘covid 19’ OR ‘sars coronavirus’/exp OR ‘sars cov 2’ OR ‘coronavirus disease 2019’/exp OR coronavirus OR ‘2019 ncov infection’ OR ‘2019 ncov disease’

#2 ‘pharmacist’/exp OR pharmacist OR pharmacists OR ‘community pharmacist’/exp OR ‘community pharmacists’ OR ‘clinical pharmacist’/exp OR ‘clinical pharmacists’ OR ‘hospital pharmacist’/exp OR ‘hospital pharmacists’ OR ‘hospital pharmacy’/exp OR ‘hospital pharmacy’ OR ‘clinical pharmacy’/exp OR ‘clinical pharmacy’ OR ‘pharmacy (shop)’/exp OR ‘community pharmacy’ OR pharmacy OR pharmacies OR ‘pharmaceutical care’/exp OR ‘pharmaceutical care’ OR ‘medication therapy management’/exp OR ‘medication therapy management’ OR ‘drug therapy management’ OR ‘medication management’

#1 AND #2

Filter: publication date from 2019/12/01

### C. Scopus

#1 TITLE-ABS-KEY ((covid 19) OR (sars AND cov 2) OR coronavirus OR (2019 ncov AND infection) OR (2019 ncov AND disease))

#2 TITLE-ABS-KEY (pharmacist OR pharmacists OR (community AND pharmacists) OR (clinical AND pharmacists) OR (hospital AND pharmacists) OR (hospital AND pharmacy) OR (clinical AND pharmacy) OR (community AND pharmacy) OR pharmacy OR pharmacies OR (pharmaceutical AND care) OR (medication AND therapy AND management) OR (drug AND therapy AND management) OR (medication AND management))

#1 AND #2

Filter: publication date from 2019/01/01

### D. LILACS

(tw:(covid-19 OR coronavirus OR “sars-Cov-2” OR “2019 ncov infection” OR “2019 ncov disease”)) AND (tw:(pharmacist OR pharmacists OR “community pharmacists” OR “clinical pharmacists” OR “hospital pharmacists” OR “hospital pharmacy” OR “clinical pharmacy” OR “community pharmacy” OR pharmacy OR pharmacies OR “pharmaceutical care” OR “medication therapy management” OR “drug therapy management” OR “medication management”))

Filter: publication date from 2019/01/01

### E. DOAJ

covid-19 AND pharmacy

## Appendix 2.

List of excluded studies.

| Reason for exclusion | Authors, year | Title | Reference |
| --- | --- | --- | --- |
| Not described activities and-or services adopted by the pharmacist | Shubha B, Amanuel K | Additional clinical pharmacists roles during COVID-19 | J Am Coll Clin Pharm.<br>2020 Apr 12 |
|  | Vouri SM, et al. | An evaluation of co-use of chloroquine or hydroxychloroquine plus azithromycin on cardiac outcomes: A pharmacoepidemiological study to inform use during the COVID19 pandemic | Res Social Adm Pharm.<br>2020 Apr 30 |
|  | Brown JD, et al. | Cannabidiol as Prophylaxis for SARS-CoV-2 and COVID-19? Unfounded Claims Versus Potential Risks of Medications During the Pandemic | Res Social Adm Pharm.<br>2020 Mar 31;S1551-7411(20)30300-4. |
|  | Carico Jr RR, et al. | Community Pharmacists and Communication in the Time of COVID-19: Applying the Health Belief Model | Res Social Adm Pharm.<br>2020 Mar 26;S1551-7411(20)30293-X |
|  | Hedima EW,<br>Adeyemi MS,<br>Ikunaiye NY | Community Pharmacists: On the frontline of health service against COVID-19 in LMICs | Res Social Adm Pharm.<br>2020 Apr 17 |
|  | Khan Z, et al. | Coronavirus outbreaks: prevention and management recommendations | Drugs Ther Perspect.<br>2020;36:215–217. |
|  | Fowler P | COVID-19: A chance to be our best | J Pharm Pract Res 2020<br>April 30 |
|  | Ailabouni NJ,<br>et al. | COVID-19 Pandemic: Considerations for Safe Medication Use in Older Adults with Multimorbidity and Polypharmacy | J Gerontol A.<br>2020;26(6):1-15 |
|  | Iacobucci G | Covid-19: Allow pharmacists to dispense controlled drugs without prescription, urge specialists | BMJ. 2020 May<br>7;369:m1868. |
|  | Adunlin G,<br>Murphy PZ,<br>Manis M. | COVID-19: How Can Rural Community Pharmacies Respond to the Outbreak? | J Rural Health. 2020 Apr<br>11. |
|  | Zanon D, et al. | Data on the stability of darunavir/cobicistat suspension after tablet manipulation | Data Brief.<br>2020;30;105552 |
|  | Métayer R | Dispositif d'urgence en situation de crise sanitaire: application d'un régime dérogatoire au pharmacien d'officine | Médecine & Droit. 2020 Apr 17 |
|  | EHAP | EHAP launches COVID-19 resource centre for hospital pharmacist | https://www.eahp.eu/hp-practice/hospital-pharmacy/eahp-covid-19-resource-centre |
|  | Meghana A, et al. | Emergency preparedness and response (EP&R) by pharmacy professionals in India: Lessons from the COVID-19 pandemic and the way forward | Res Social Adm Pharm. 2020 Apr 25 |
|  | Dawoud D | Emerging from the other end: Key measures for a successful COVID-19 lockdown exit strategy and the potential contribution of pharmacists. | Res Social Adm Pharm. 2020 May 13 |
|  | Wiffen P. | Houston we have a problem: coronavirus! | Eur J Hosp Pharm. 2020 Mar;27(2):59 |
|  | Amariles P, et al | How to link patients with suspicious COVID-19 to health system from the community pharmacies? A route proposal | Res Social Adm Pharm. 2020 Mar 23;S1551-7411(20)30248-5 |
|  | Kretchy IA, Asiedu-Danso M, Kretchy JP | Medication management and adherence during the COVID-19 pandemic: Perspectives and experiences from low-and middle-income countries | Res Social Adm Pharm. 2020 Apr 15 |
|  | Cadogan CA, Hughes CM | On the frontline against COVID-19: Community pharmacists' contribution during a public health crisis | Res Social Adm Pharm. 2020 Mar 31;S1551-7411(20)30292-8 |
|  | Alderman C. | On the Pharmacy Radar: COViD-19 and Older People | Sr Care Pharm. 2020;35(5):190-194. |
|  | Green TC, Bratberg J, Finnell DS. | Opioid use disorder and the COVID 19 pandemic: A call to sustain regulatory easements and further expand access to treatment | Subst Abus. 2020;41(2):147-149. |
|  | Ha DR, et al. | Outpatient hydroxychloroquine prescribing at a large academic health system during the COVID-19 pandemic | Infect Control Hosp Epidemiol. 2020 May 15 |
|  | Bahlol M,<br>Deweyc RS | Pandemic preparedness of community pharmacies for COVID-19 | Res Social Adm Pharm.<br>2020 May 11 |
|  | Bukhari N, et<br>al. | Pharmacists at the frontline beating the COVID-19 pandemic | J Pharm Policy Pract.<br>2020;13(8) |
|  | Aruru M,<br>Truong HA,<br>Clark S. | Pharmacy Emergency Preparedness and Response (PEPR): a proposed framework for expanding pharmacy professionals' roles and contributions to emergency preparedness and response during the COVID-19 pandemic and beyond | Res Social Adm Pharm.<br>2020 Apr 10 |
|  | Alderman C. | Pharmacy Services and the Novel Coronavirus | Sr Care Pharm.<br>2020;35(4):146-147 |
|  | Mukhatar S. | Preparedness and proactive infection control measures of Pakistan during COVID-19 pandemic outbreak | Res Social Adm Pharm.<br>2020 Apr 11;S1551-7411(20)30373-9. |
|  | Stergachis A | Preparing pharmacy for the surge of patients with COVID-19: Lessons from China | J Am Pharm Assoc. 2020<br>Apr 10;S1544-3191(20)30152-7. |
|  | Zheng SQ, et al. | Recommendations and guidance for providing pharmaceutical care services during COVID-19 pandemic: A China perspective | Res Social Adm Pharm. 2020 Mar 26;S1551-7411(20)30284-9. |
|  | Bhat S, et al. | Roles of Clinical Pharmacists in Caring for Patients with Inflammatory Bowel Disease during COVID-19 | Gastroenterology. 2020 May 13 |
|  | Gross AE. | Roles of the clinical pharmacist during the COVID-19 pandemic | J Am Coll Clin Pharm. 2020 Mar 30 |
|  | Al-Quteimat OM, Amer AM. | SARS-CoV-2 outbreak: How can pharmacists help? | Res Social Adm Pharm. 2020 Mar 26;S1551-7411(20)30238-2. |
|  | Hasan SS, Kow CS, Zaidi STR | Social distancing and the use of PPE by community pharmacy personnel: Does evidence support these measures? | Res Social Adm Pharm. 2020 May 1;S1551-7411(20)30490-3. |
|  | Watson KE, et al. | The evolution of pharmacists' roles in disasters, from logistics to assessing and prescribing | Can Pharm J (Ott). 2020;153(3):129-131. |
|  | Vinci DL,<br>Polidori C,<br>Polidori P | The healthcare and pharmaceutical vulnerability emerging from the new Coronavirus outbreak | Eur J Hosp Pharm.<br>2020;27(3):129-130. |
|  | Chan AHY, et al. | Together we unite: the role of the Commonwealth in achieving universal health coverage through pharmaceutical care amidst the COVID-19 pandemic | J Pharm Policy Pract.<br>2020;13:13. |
|  | Ziegler A. | Wenn keiner liefern kann | Dtsch Apoth Ztg.<br>2020;32(10):1-7 |
|  | Erku DA, et al. | When fear and misinformation go viral: Pharmacists' role in deterring medication misinformation during the 'infodemic' surrounding COVID-19. | Res Social Adm Pharm.<br>2020 May 1 |
| Reviews, recommendations, and-or guidelines | Mahmoudjafari Z, et al. | American Society for Transplantation and Cellular Therapy Pharmacy Special Interest Group Position Statement on Pharmacy Practice Management and Clinical Management for COVID-19 in Hematopoietic Cell Transplantation and Cellular Therapy Patients in the United States | Biol Blood Marrow Transplant. 2020 Jun;26(6):1043-1049. |
|  | Li H, et al. | Fighting Against COVID-19: Innovative Strategies for Clinical Pharmacists | Res Social Adm Pharm. 2020 Apr 6;S1551-7411(20)30328-4. |
|  | Song Z, et al. | Hospital pharmacists' pharmaceutical care for hospitalized patients with COVID-19: Recommendations and guidance from clinical experience | Res Social Adm Pharm. 2020 Apr 3. doi: 10.1016/j.sapharm.2020.03.027 |
|  | Liu S et al. | Providing pharmacy services during the coronavirus pandemic | Int J Clin Pharm. 2020;42:299-304. |
|  | Lin ZJ, Zhang B | Strategy of pharmaceutical care services for clinical Chinese pharmacists in coronavirus disease 2019 (COVID-19) | Zhongguo Zhong Yao Za Zhi. 2020;45(6):1259-1262. |
| Pharmacotherapeutic options | Jiang S, et al. | Pharmaceutical care for severe and critically ill patients with COVID-19 | J Zhejiang Univ. 2020;49(2): 158-169 |
|  | Sun X, et al. | Pharmaceutical care of chloroquine phosphate in elderly patients with coronavirus pneumonia (COVID-19) | Aging Med. 2020. April 6. |
|  | Wang Y, Zhu LQ. | Pharmaceutical care recommendations for antiviral treatments in children with coronavirus disease 2019 | World J Pediatr. 2020 Mar 12;1-4 |
|  | Zhang S, et al. | Rational Use of Tocilizumab in the Treatment of Novel Coronavirus Pneumonia | Clin Drug Investig. 2020 Apr 26 : 1–8. |
| Not related the pharmacist professional | Stevens RW, Estes L, Rivera C. | Practical Implementation of COVID-19 Patient Flags into an Antimicrobial Stewardship Program's Prospective Review | Infect Control Hosp Epidemiol. 2020 Apr 15 |
| Involved graduate students | Basheti IA, et al. | Pharmacists' readiness to deal with the coronavirus pandemic: Assessing awareness and perception of roles | Res Social Adm Pharm. 2020 Apr 23;S1551-7411(20)30418-6 |
| Interview | Dedy J. | Schwere COVID-19-Fälle gemeinsam versorgen | Dtsch Apoth Ztg. 2020;36(17). |
| No access | Xie F, et al. | Preventive strategy for tumor patients during COVID-19 pandemic | J. Pract. Oncol. 2020;35(2):123-126. |

